# Heterogeneous distribution of tau pathology in the behavioral variant of Alzheimer’s disease

**DOI:** 10.1101/2020.09.18.20188276

**Authors:** Ellen H. Singleton, Oskar Hansson, Yolande A. M. Pijnenburg, Renaud La Joie, William G. Mantyh, Pontus Tideman, Erik Stomrud, Antoine Leuzy, Maurits Johansson, Olof Strandberg, Ruben Smith, Evi Berendrecht, Bruce Miller, Leonardo Iaccarino, Lauren Edwards, Amelia Storm, Emma Wolters, Emma M. Coomans, Denise Visser, Sandeep S.V. Golla, Hayel Tuncel, Femke Bouwman, John van Swieten, Janne M. Papma, Bart van Berckel, Philip Scheltens, Anke A. Dijkstra, Gil Rabinovici, Rik Ossenkoppele

## Abstract

**Objective:** The clinical phenotype of the rare behavioral variant of Alzheimer’s disease (bvAD) is insufficiently understood. Given the strong clinico-anatomical correlations of tau pathology in AD, we investigated the distribution of tau deposits in bvAD, *in-vivo* and *ex-vivo*, using PET and postmortem examination.

**Methods:** For the tau PET study, seven amyloid-β positive bvAD patients underwent [^18^F]flortaucipir or [^18^F]RO948 PET. We converted tau PET uptake values into standardized (W-)scores, by adjusting for age, sex and MMSE in a “typical” memory-predominant AD (n=205) group. W-scores were computed within entorhinal, temporoparietal, medial and lateral prefrontal, insular and whole-brain regions-of-interest, frontal-to-entorhinal and frontal-to-parietal ratios and within intrinsic functional connectivity network templates. For the postmortem study, the percentage of AT8 (tau)-positive area in hippocampus CA1, temporal, parietal, frontal and insular cortices were compared between autopsy-confirmed bvAD (n=8) and typical AD (n=7) patients.

**Results:** Regional W-scores ≥1.96 (corresponding to p<0.05) were observed in three cases, i.e. case #5: medial prefrontal cortex (W=2.13) and anterior default mode network (W=3.79), case #2: lateral prefrontal cortex (W=2.79) and salience network (W=2.77), and case #7: frontal-to-entorhinal ratio (W=2.04). The remaining four cases fell within the normal distributions of the typical AD group. Postmortem AT8 staining indicated no regional differences in phosphorylated tau levels between bvAD and typical AD (all p>0.05).

**Conclusion:** Both *in-vivo* and *ex-vivo*, bvAD patients showed heterogeneous patterns of tau pathology. Since key regions involved in behavioral regulation were not consistently disproportionally affected by tau pathology, other factors are more likely driving the clinical phenotype in bvAD.

## INTRODUCTION

Individuals with the behavioral variant of Alzheimer’s disease (bvAD, previously referred to as “frontal AD”) experience early prominent behavioral symptoms and personality changes, such as disinhibition, compulsive behaviors and loss of empathy^1, 2^ These individuals are clinically reminiscent of behavioral variant frontotemporal dementia (bvFTD), but have AD as the primary pathology and resemble patients with “typical” AD (tAD) neuroanatomically, as atrophy and hypometabolic patterns in bvAD predominantly occur in temporoparietal regions^1, 3^. Imaging and pathological investigations (mostly case reports or small cohort studies based on the low prevalence of this phenotype^1^) have provided mixed results regarding the involvement of the frontal cortex in bvAD^4-8^. This apparent clinico-anatomical dissociation indicates the need for a better understanding of the neurobiological factors underlying the bvAD phenotype. To that end, it is crucial to study the distribution of tau deposition in bvAD, as this central neuropathological hallmark of AD is closely related to type and severity of cognitive symptoms^9, 10^ and precedes and predicts patterns of neurodegeneration detected by MRI and [^18^F]FDG PET^11, 12^ To date, only two bvAD case studies with tau PET have been reported, one showing a posterior pattern of tau pathology^9^ and the other one a more diffuse uptake pattern including both temporoparietal and frontal regions^13^. In this study, we had three objectives: to investigate the regional distribution of tau pathology in bvAD i) *in-vivo* using tau PET and ii) *ex-vivo* using postmortem examination, and iii) to assess the relationship between tau PET patterns and behavioral symptoms in typical AD.

## METHODS

### Participants

For the tau PET study, we included seven patients clinically diagnosed with bvAD from the Amsterdam Dementia Cohort (ADC, the Netherlands, n=2), the University of California San Francisco (UCSF, United States, n=3) Alzheimer Disease Research Center and the Swedish BioFINDER study (http://www.biofinder.se; Sweden, n=2). In the absence of formal clinical consensus criteria for bvAD we used our previously established procedure^1^. First, among participants with available tau PET, we selected those with a clinical diagnosis of AD dementia^14^ or mild cognitive impairment (MCI)^15^. From this selection we included only patients who were on the AD pathological continuum according to the NIA-AA research criteria^11^ of amyloid-β positivity based on PET or cerebrospinal fluid (CSF). Third, we performed extensive chart reviews (by R.O.) and included only participants fulfilling ≥2 of 6 core clinical criteria for bvFTD^16^, consisting of apathy, loss of empathy, disinhibition, compulsive behaviors, hyperorality and dysexecutive functioning. This ensured the inclusion of patients with robust and clinically prominent “bvFTD-like” symptoms, and was based on our previous finding that 75% of bvAD patients showed ≥2 bvFTD clinical symptoms, and bvAD patients generally showed a slightly milder behavioral profile than bvFTD patients^1^. Note that we excluded participants with dysexecutive AD^17^ if they did not meet any of the remaining five bvFTD criteria, in order to selectively study the abovementioned core behavioral features in AD. We compared the participants with bvAD to participants with typical AD from all centers (ADC, n=55; UCSF, n=60; BioFINDER, n=90), consisting of Aβ-positive AD dementia and mild cognitive impairment (MCI) participants who had undergone tau PET. Participants meeting diagnostic criteria for posterior cortical atrophy or the logopenic variant of primary progressive aphasia were excluded from this group. In addition, patients with known autosomal dominant mutations for AD or FTD were excluded. A clinical description of the bvAD cases can be found in e-table 1. For the postmortem study, eight patients clinically diagnosed with bvAD who donated their brains to the Netherlands Brain Bank were compared to seven participants with typical AD. These diagnoses were established retrospectively based on antemortem clinical diagnosis of “frontal variant of AD”, bvFTD or a differential diagnosis of bvFTD vs AD^1^. All bvAD and typical AD patients had a primary neuropathological diagnosis of AD. For the assessment of relationships between tau PET patterns and behavioral symptoms in typical AD, we included patients from the typical AD groups in the tau PET study that had behavioral measures available (ADC, n=28; UCSF, n=48; BioFINDER; n=97). Informed consent was obtained from all participants and local institutional review boards for human research approved this study.

**Table 1.** Characteristics of participants with the behavioral variant of Alzheimer’s disease and participants with typical Alzheimer’s disease in the PET study.

|  | **ADC** |  | **UCSF** |  | **BioFINDER** |  |
| --- | --- | --- | --- | --- | --- | --- |
|  | bvAD | Typical AD | bvAD | Typical AD | bvAD | Typical AD |
| N | 2 | 55 | 3 | 60 | 2 | 90 |
| Age (mean) | 68 | 65.7 (7.7) | 66 | 64.5 (8.8) | 75 | 73.1 (6.6) |
| Sex (%male) | 100% | 49% | 100% | 42% | 50% | 54% |
| Education (y) | 13.0 | 12.2 (3.1)<sup>a</sup> | 17.0 | 17.3 (3.1)<sup>c</sup> | 8.0 | 12.2 (4.9)<sup>f</sup> |
| MMSE | 20 | 23.2 (3.9) | 21 | 22.1 (6.5)<sup>d</sup> | 25 | 20.2 (4.1)<sup>g</sup> |
| APOE ε4 pos | 0% | 79%<sup>b</sup> | 50% | 58%<sup>e</sup> | 100% | 73%<sup>h</sup> |

<sup>a</sup>n=51 for education ADC, <sup>b</sup>n=47 for APOE ε4 ADC, <sup>c</sup>n=48 for education UCSF, <sup>d</sup>n=50 for MMSE UCSF, <sup>e</sup>n=43
for APOE ε4 UCSF, <sup>f</sup>n=86 for education BioFINDER, <sup>g</sup>n=89 for MMSE BioFINDER, <sup>h</sup>n=88 for APOE ε4
BioFINDER. Error margins were not included for bvAD patients for privacy reasons.

### Standard protocol approvals, registrations and patient consents

Informed consent was obtained from all subjects or their assigned surrogate decision-makers, and the study was approved by the Amsterdam University Medical Center, Memory and Aging Center Clinic at the University of California San Francisco and the Memory Clinic Skåne University Hospital institutional human research review boards.

### Tau PET in bvAD compared to typical AD

PET scanning was performed using the tau tracers [^18^F]flortaucipir (ADC, UCSF) and [^18^F]RO948 (BioFINDER). Image acquisition and processing for each center have been described previously^9, 18, 19^ and are summarized in e-table 2. Briefly, we generated standardized uptake value ratios (SUVR) for the interval between 80-100 ([^18^F]flortaucipir) or 70-90 ([^18^F]RO948) minutes post-injection using (inferior) cerebellar gray cortex as the reference region. We then computed native space derived mean SUVR values in the following (composite) regions-of-interests (ROIs) representing a mix of AD and bvFTD vulnerable regions: entorhinal, temporoparietal, frontal, and insular cortices, and whole cortex. To examine the relative tau burden in frontal regions compared to classical AD regions, we additionally computed frontal-to-entorhinal and frontal-to-parietal ratios. A detailed composition of each ROI is shown in e-table 3. Furthermore, mean SUVR values were extracted from four functional connectivity network templates in MNI space implicated in AD and bvFTD, including the executive control network, salience network, anterior default mode network and posterior default mode network^20^. For each ROI we computed W-scores reflecting standardized individual differences between the observed and predicted SUVR based on the typical AD distribution, adjusted for age, sex and MMSE score (i.e. W=(observed SUVR-predicted SUVR)/SDresiduals)). Note that the limited sample size and differences in tau PET acquisition across cohorts did not allow group-wise statistical comparisons, hence results are described as the W-score in individual bvAD patients relative to the normal distribution across the typical AD group (i.e. W scores ≥1.96, corresponding to p<0.05). For visual purposes, the co-registered T1-weighted MRI scans were warped to Montreal Neurological Institute (MNI152) space, and these transformation matrixes were applied to warp native space SUVR images to MNI space. The normalized PET images were then smoothed using an 8-mm Gaussian kernel. The tau PET images of individual bvAD patients were visually compared to an average SUVR image for the (cohort-specific) typical AD groups.

**Table 2.** Behavioral symptoms and regional tau PET deposition in individual bvAD cases versus typical AD patients presented as a group.

|  | **ADC** |  |  | **UCSF** |  |  | **BioFINDER** |  |  |  |
| --- | --- | --- | --- | --- | --- | --- | --- | --- | --- | --- |
|  | bvAD1 | bvAD2 | Typical AD | bvAD3 | bvAD4 | bvAD5 | Typical AD | bvAD6 | bvAD7 | Typical AD |
| **bvFTD criteria** |  |  |  |  |  |  |  |  |  |  |
| Disinhibition | Y | N |  | Y | Y | N |  | Y | Y |  |
| Apathy | Y | Y |  | Y | N | Y |  | Y | Y |  |
| Loss of empathy | Y | N |  | N | N | Y |  | Y | N |  |
| Compulsiveness | N | Y |  | Y | Y | N |  | N | N |  |
| Hyperorality | Y | Y |  | N | N | N |  | N | Y |  |
| Dysexecutive | N | N |  | N | N | N |  | Y | N |  |
| n | 4 | 3 |  | 3 | 2 | 2 |  | 4 | 3 |  |
| **Tau SUVR** |  |  |  |  |  |  |  |  |  |  |
| Entorhinal | 1.27 | 1.81 | 1.49 (0.25) | 1.53 | 1.49 | 1.89 | 1.73 (0.35) | 1.59 | 1.91 | 1.98 (0.42) |
| Temporoparietal | 1.56 | 1.80 | 1.58 (0.40) | 1.67 | 1.74 | 1.76 | 1.94 (0.56) | 1.40 | 2.28 | 1.84 (0.61) |
| Medial frontal | 1.45 | 1.53 | 1.25 (0.28) | 1.39 | 1.50 | 2.05 | 1.48 (0.35) | 1.15 | 1.77 | 1.38 (0.54) |
| Lateral frontal | 1.58 | 2.21 | 1.39 (0.38) | 1.43 | 1.69 | 1.88 | 1.68 (0.50) | 1.15 | 2.28 | 1.46 (0.63) |
| Insula | 1.43 | 1.24 | 1.22 (0.15) | 1.12 | 1.14 | 1.35 | 1.22 (0.14) | 1.06 | 1.12 | 1.19 (0.21) |
| Mean cortical | 1.51 | 1.62 | 1.44 (0.30) | 1.53 | 1.58 | 1.82 | 1.72 (0.43) | 1.30 | 2.10 | 1.66 (0.51) |
| Frontal:Entorhinal | 1.19 | 0.99 | 0.89 (0.21) | 0.92 | 1.07 | 1.04 | 0.92 (0.21) | 0.78 | 1.16 | 0.78 (0.22) |
| Frontal:Parietal | 1.04 | 0.92 | 0.84 (0.14) | 0.79 | 0.89 | 1.18 | 0.83 (0.16) | 0.97 | 0.96 | 0.90 (0.20) |

**Table 3.** Characteristics of participants with behavioral variant of Alzheimer’s disease and typical Alzheimer’s disease in the postmortem study.

|  | **bvAD** | **Typical AD** |
| --- | --- | --- |
| n | 8 | 7 |
| Age (mean) | 67 | 69.1 (3.3) |
| Sex (% male) | 50% | 29% |
| Disease duration (months) | 75 | 55.0 (39.9) |
| % AD primary Dx neuropathology | 87.5% | 100% |
| % A3B3C3 score | 87.5% | 100% |
| Brain weight (grams) | 1093.5 | 982.1 (192.7) |
| PMI | 6.5 | 4.9 (1.1) |
| % CAA present | 62.5% | 71.4% |
| % CVD present | 50.0% | 57.1% |
| % LB present | 75.0% | 57.1% |

*bvAD*=behavioral variant Alzheimer’s disease, *M*=male, *F*=female, *MMSE*=Mini Mental State Examination,
*ABC score*=Amyloid, Braak, CERAD criteria, *AD*=Alzheimer’s disease, *DLB*=dementia with Lewy bodies,
*PMI*=post-mortem interval in hours, *n/a or -*=not available, *CAA*=cerebral amyloid angiopathy, *CVD*=cerebral
vascular disease, *LB*=Lewy bodies, *TDP*=TAR DNA binding protein, *ARTAG*=Aging-related tau astroglipathy.
Error margins were not included for bvAD patients for privacy reasons.

### Associations between tau PET patterns and age in bvAD relative to typical AD

We then examined the influence of age-of-onset on the involvement of the frontal regions in bvAD, as younger age has previously been linked to great tau pathology across the neocortex^21^. Therefore, the associations between age and tau PET uptake in medial prefrontal, lateral prefrontal, salience network and anterior default mode network regions were plotted and the tau PET SUVR’s of the bvAD cases were studied relative to the distribution the typical AD age groups.

### Postmortem investigation of tau pathology in bvAD compared to typical AD

Immunohistochemistry was performed with antibodies against phosphorylated tau using AT8 (AT8 antibody, 1:800 dilution, ThermoFisher, Waltham, USA) on 8 μm thick representative sections of the anterior cingulate cortex, hippocampus CA1, caudate nucleus, entorhinal cortex, frontal pole, frontoinsula, putamen, subiculum and thalamus of the right hemisphere. A detailed description of the procedures can be found in e-table 3. The presence of chromogen 3.3’-diaminobenzidine (DAB: K5007; DAKO) staining was quantified using the color threshold plugin in ImageJ (version 1.52u; NIH), where the threshold was set to include tangles and threads. Of each region, two images were taken and the outcome measurement was the average percentage of DAB-stained pixels per brain region. Systematic staining was performed for Aβ42, α-synuclein and 3R and 4R tau, but not for TDP-43 (only if there was a clear indication). Between group differences in percentage of tau pathology brain region were assessed using Mann Whitney U tests, adjusting for age and sex.

### Associations between regional tau PET uptake and behavioral symptoms in typical AD

Finally, we tested whether regional tau pathology in typical AD patients was associated with behavioral disturbances within the realm of bvFTD clinical symptoms. At the ADC, the apathy, disinhibition and eating abnormalities domains (frequency(1-4)*severity(0-3)) of the Neuropsychiatric Inventory (NPI)^22^ were averaged to create a bvFTD composite score. For BioFINDER, the motivation, impulse control, and social domains (severity score (1-3)) of the Mild Behavioral Impairment-Checklist (MBI-C)^23^ were averaged (average score=(total score on apathy+total score on impulse control+total score on social domain)/3). As the subscales of the MBI-C differ in the number of items, analyses were repeated using a weighted bvFTD composite score (weighted score=scores on each item of each relevant domain/total amount of items). In a sensitivity analysis, associations between regional tau PET uptake and separate subdomain scores were studies for both the ADC and BioFINDER cohort. At UCSF, scores on an Affect Naming task^24^ assessing emotion recognition, a subcomponent of social cognition, were used. These three measures across the three centers will be termed “bvFTD measures” throughout this manuscript. High scores indicate more behavioral disturbances. The behavioral questionnaires were performed half a year on average before or after the tau PET acquisitions. E-table 4 includes a detailed description of the tests and procedures. The same tau PET ROIs and ratios described above were used for this analysis. Spearman rank correlations were used between SUVR in tau regions/ratios and the bvFTD measures, adjusting for age and sex. We used R v4.0.2 (https://www.R-project.org/) for statistical analyses^25^. A p-value below 0.05 was considered significant.

### Data availability statement

Anonymized data used in the present study may be available upon request to the corresponding author.

## RESULTS

### Demographic characteristics participants with bvAD

Demographic and clinical characteristics of the participants are presented in tables 1, 2 and 3. In the tau PET study, 6/7 (85.7%) bvAD cases were male, while 48.3% of typical AD patients were male. Age ranged from 59 to 80 in the bvAD cases (mean: 69.1±8.4), compared to a mean age of 67.8±7.7 in the typical AD groups. MMSE ranged between 17 and 26 in bvAD cases (mean: 21.7±2.8), with average MMSE scores of 21.8±4.8 in the typical AD cases (table 1). 3/7 bvAD cases were *APOE* ε4 positive, ε/7 *APOE* ε3 homozygote, and *APOE* genotype was missing for 1 bvAD case, while *APOE* ε4 positivity was found in 70% of the typical AD cases. Presence of bvFTD symptoms (maximum is 6) ranged from 2 to 6 in bvAD cases, with apathy as the most prevalent symptom (n=6), followed by disinhibition (n=5), loss of empathy, compulsiveness and hyperorality (all n=3), and dysexecutive profile (n=1). In the postmortem study, 4/8 (50.0%) bvAD cases were male versus 3/7 (42.9%) in the typical AD group, and the mean age at death was 66.6±6.0 in the bvAD group versus a mean age of 69.1±3.3 in the typical AD group. Disease duration was slightly longer in bvAD cases (6.3±3.6 years) compared to typical AD cases (4.6±3.3 years).

### Tau PET in bvAD compared to typical AD

Figure 1 shows the tau PET patterns for all individual bvAD cases relative to an average tau PET image for the whole typical AD group per cohort. Visual assessment indicated that 3/7 bvAD cases (#2, #5, and #7) showed prominent frontal involvement in addition to substantial temporoparietal uptake. Among these cases, case #5 showed strongly elevated uptake in the medial prefrontal cortex, while #2 and #7 showed predominant lateral frontal uptake. One case (#4) showed some uptake in the lateral frontal cortex, but the medial parietal cortex was clearly the most affected brain region. 2/7 cases (#1 and #6) had a lateral temporal predominant uptake pattern with very limited frontal involvement. One case (#3) showed a classical AD-like temporoparietal uptake pattern with minimal tracer retention in the frontal cortex. The heterogeneity in tau patterns across bvAD patients was confirmed by quantitative ROI analyses, showing W-scores ≥1.96 only in one case in the medial prefrontal (#5, W=2.13) and lateral prefrontal (#2, W=2.79) regions and in one case in the ratio frontal-to-entorhinal tau (#7, W=2.04; e-table 5 & figure 2). All ROIs and ratios in the remaining four cases fell within the normal distribution of the typical AD group. Regarding tau uptake within functional connectivity network templates, W-scores ≥1.96 were found in one case (#2, W=2.77) in the salience network and another case (#5, W=3.79) in the anterior default mode network (e-table 6, e-table 7 & figure 2). All network W-scores in the remaining five cases fell within the normal distribution of the typical AD group.

**Figure 1.**
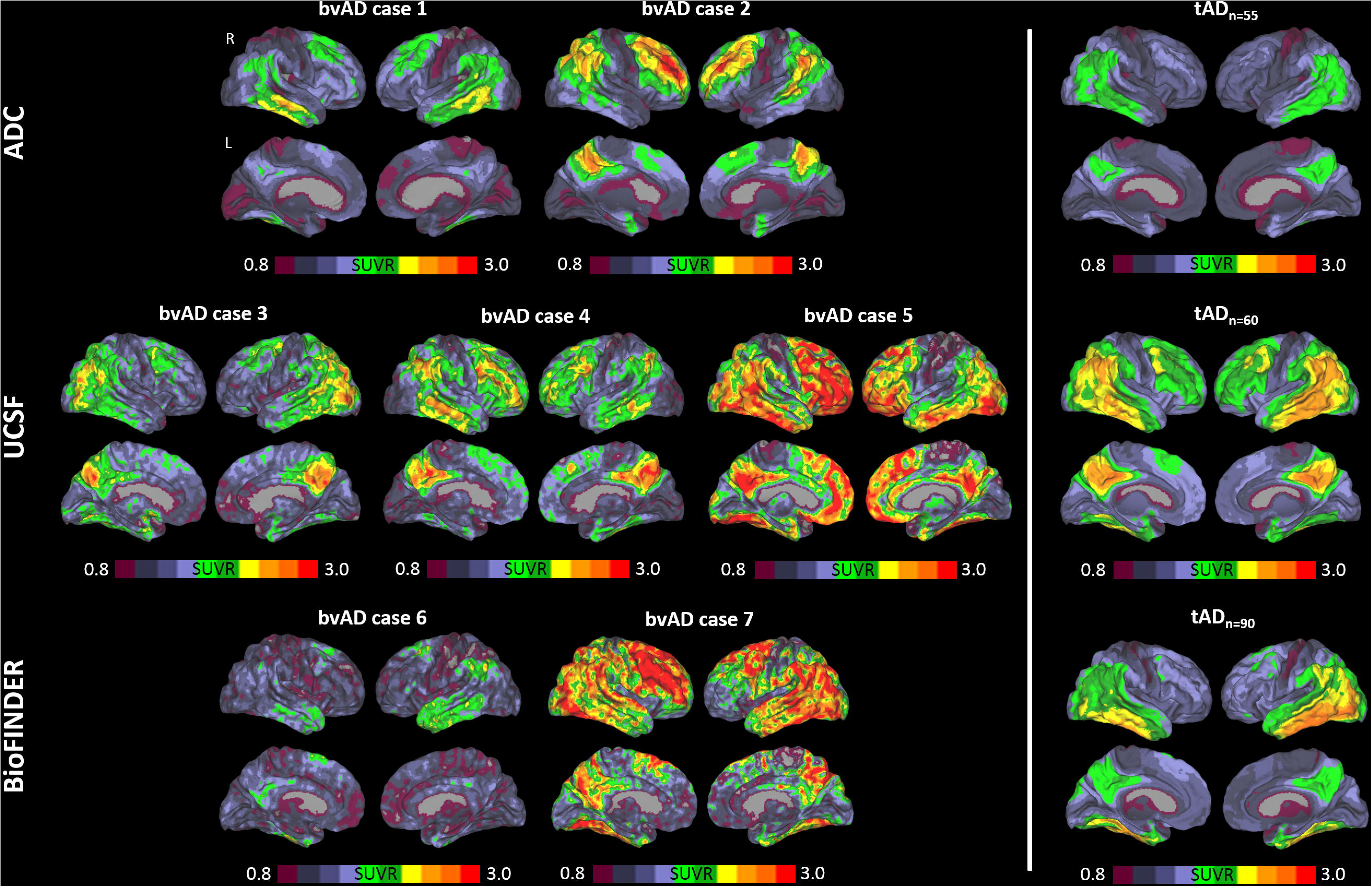
Distribution of tau pathology across the brain of participants with the behavioral variant of Alzheimer’s disease (bvAD, displayed individually) versus participants with the typical Alzheimer’s disease (tAD, displayed as the average of the group). *SUVR*=standardized uptake value ratio, *ADC*=Amsterdam Dementia Cohort, *UCSF* = University of California San Francisco.

### Associations between tau PET patterns and age in bvAD relative to typical AD

Among the three early-onset (<65 years) bvAD cases, (lateral) frontal tau PET uptake was evident in case #2, moderate in case #4 and limited in case #3 (figure 1). The late-onset bvAD cases were characterized by prominent frontal tau PET uptake in cases #5 and #7 and relative frontal sparing in cases #1 and #6. The heterogeneity of frontal involvement across the age span was further supported by the assessment tau PET uptake in four relevant brain regions/networks (figure 3). This analysis showed that three bvAD case (#2, #3 and #5) showed substantial higher tau PET uptake than estimated based on their age in the typical AD group (observed data exceeded the 95% confidence interval), while tau PET uptake in the remaining four bvAD cases largely overlapped with the 95% confidence interval of the typical AD group.

**Figure 2.**
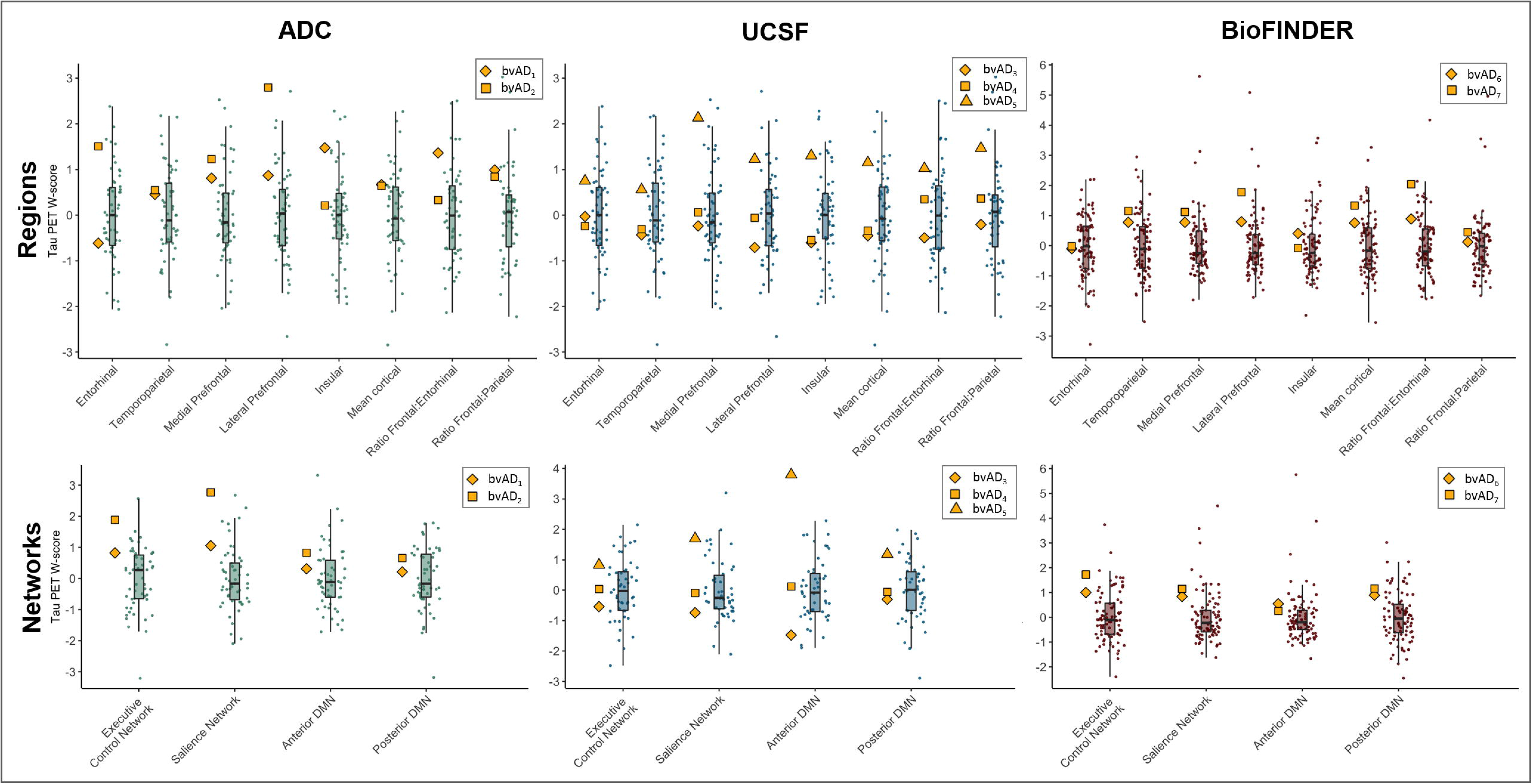
Regional tau PET retention in participants with bvAD relative to the distribution of participants with typical AD. The yellow symbols represent the individual bvAD cases and the boxplots and raincloud plots represent the distributions of the typical AD groups. *ADC*=Amsterdam Dementia Cohort, *UCSF*=University of California San Francisco, *bvAD*=behavioral variant of Alzheimer’s disease, *AD*=Alzheimer’s disease, *SUVR*=standardized uptake value ratios*, ERC*=entorhinal cortex, *TPC*=temporoparietal cortex, *FC*=frontal cortex, *IC*=insular cortex, *MC*=mean cortical uptake, *ratFE*=ratio frontal-to-entorhinal tau PET retention, *ratFP*=ratio frontal-to-parietal tau PET retention.

**Figure 3.**
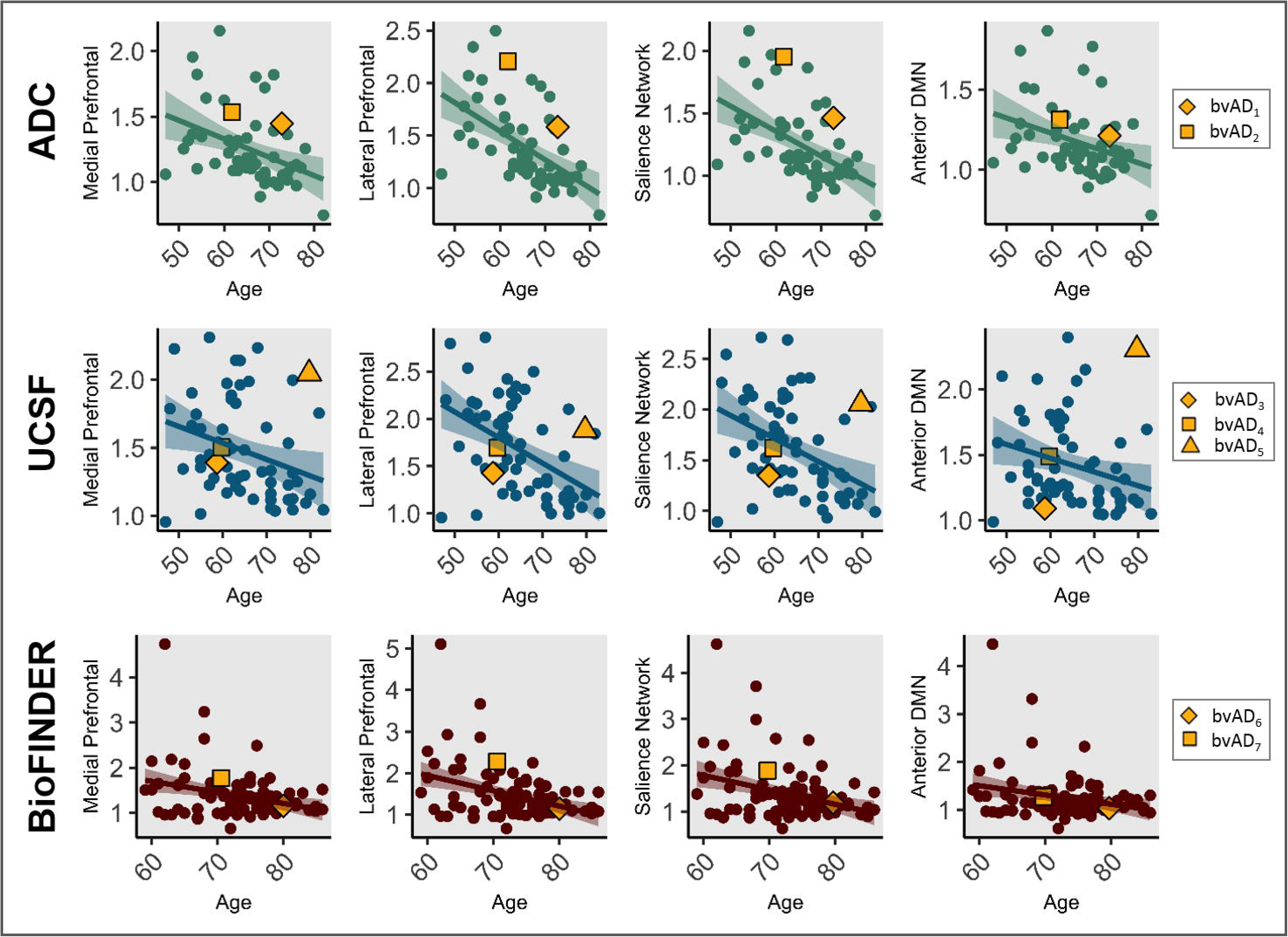
Scatterplots depicting the relationship between frontal, medial prefrontal and lateral prefrontal tau uptake and age in typical AD and bvAD across centers.

### Postmortem investigation of tau pathology in bvAD compared to typical AD

Presence of tau pathology quantified using AT8 immunohistochemistry did not show significant differences between bvAD and typical AD groups in any of the investigated brain regions (all p>0.05; figure 4 & e-table 8). One bvAD case had Lewy body disease as co-primary neuropathologic diagnosis in addition to AD. In terms of comorbid pathologies, Lewy body pathology was found in 6/8 bvAD patients versus 4/7 typical AD patients. Cerebral amyloid angiopathy was found in 5/8 bvAD cases and in 5/7 typical AD cases. Cerebral vascular disease was found in 4-5/8 bvAD cases and in 4/7 typical AD cases. Of three bvAD cases with TDP-43 staining being performed the stainings were negative, and in 5/8 cases the TDP-43 staining was not performed. One typical AD patient showed TDP-43 inclusions in the hippocampus and amygdala, reflecting LATE-NC stage 2, while 2/7 typical AD patients were negative for TDP-43 stainings and in 4/7 typical AD cases TDP-43 staining was not performed. In addition, in none of the bvAD cases the presence of 3R tau was observed in isolation.

**Figure 4.**
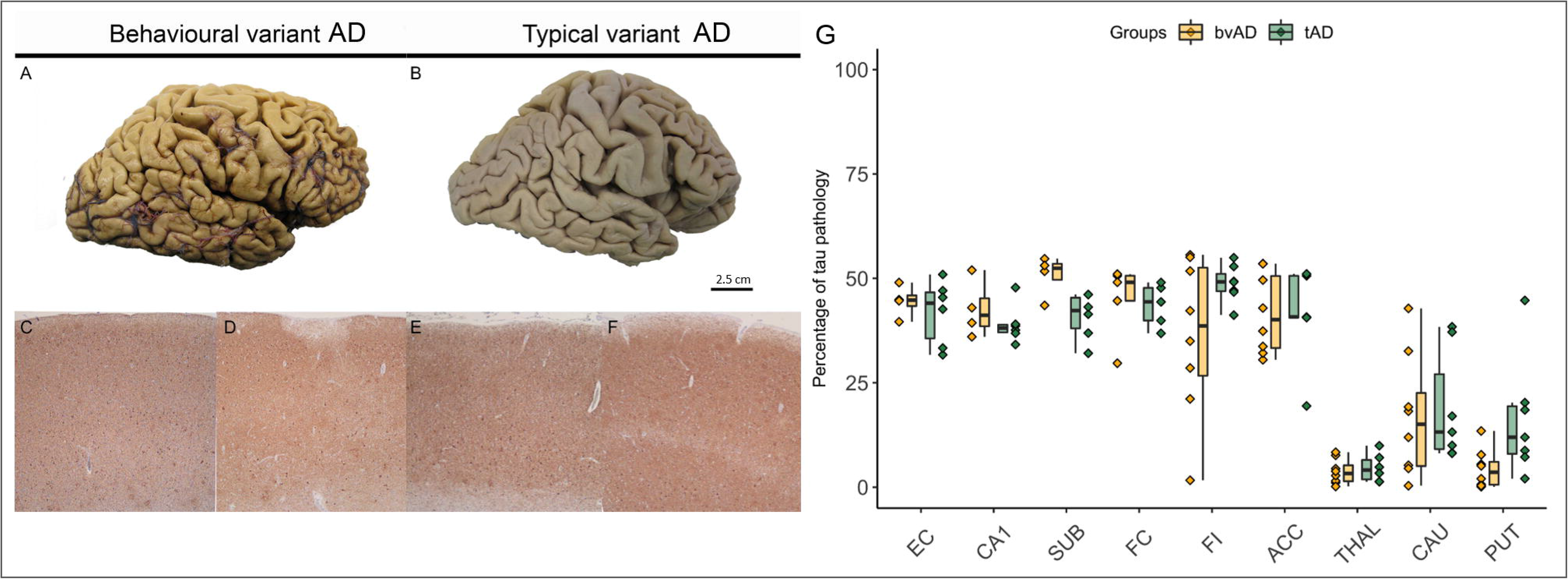
Postmortem tau immunohistochemistry in patients with bvAD and typical AD. Panel A and B show images of postmortem brain tissue of a representative case of bvAD (A) and typical AD (B), showing similar morphology. The frontal cortices are depicted in panel C and E and the entorhinal cortices are depicted in panel D and F. These images suggest that the tau burden in frontal regions in bvAD do not differ substantially from the burden in typical AD, and that the tau burdens between frontal and entorhinal cortices in both bvAD and typical AD do not differ from each other. Panel G shows the percentage of tau pathology in regions of interest in participants with bvAD and participants with typical AD, showing no significant differences between the two groups. *bvAD*=behavioral variant of Alzheimer’s disease, *AD*=Alzheimer’s disease, *EC*=Entorhinal cortex, *CA1*=Hippocampus CA1, *SUB*=Subiculum, *FC*=Frontal Cortex, *FI*=Frontoinsula, *ACC* =Anterior cingulate cortex, *THAL*=Thalamus, *CAU*=Caudate nucleus, *PUT*=Putamen.

### Associations between regional tau uptake and behavioral symptoms in typical AD

The demographic characteristics of the three typical AD cohorts (ADC, n=28; UCSF, n=48; BioFINDER, n=97) included in these analyses are shown in e-table 9. No significant relationships were found between the bvFTD clinical measures and tau deposition as measured by tau PET in any of the investigated regions (all p>0.05, figure 5), except for the frontal-to-parietal tau PET ratio in the BioFINDER cohort (rho=0.217, p=0.036). When analyses were repeated using item-weighted FTD measures, results remained the same (i.e. all p>0.05, except for the frontal-to-parietal tau PET ratio in the BioFINDER cohort, rho = 0.207, p=0.045, e-table 10). Assessment of relationships per subdomain of the NPI in the ADC cohort and the MBI in the BioFINDER cohort yielded similar results, with isolated significant results for the frontal-to-parietal ratio in the BioFINDER cohort with impulse control (rho=0.298, p=0.004) and social subdomains (rho=0.327, p=0.001), without correction for multiple comparisons, while all other associations were not significant (i.e. all p>0.05; figure e-1). This indicates that also within typical AD patients, we did not find an association between behavioral/neuropsychiatric symptoms and the degree of tau PET uptake.

**Figure 5.**
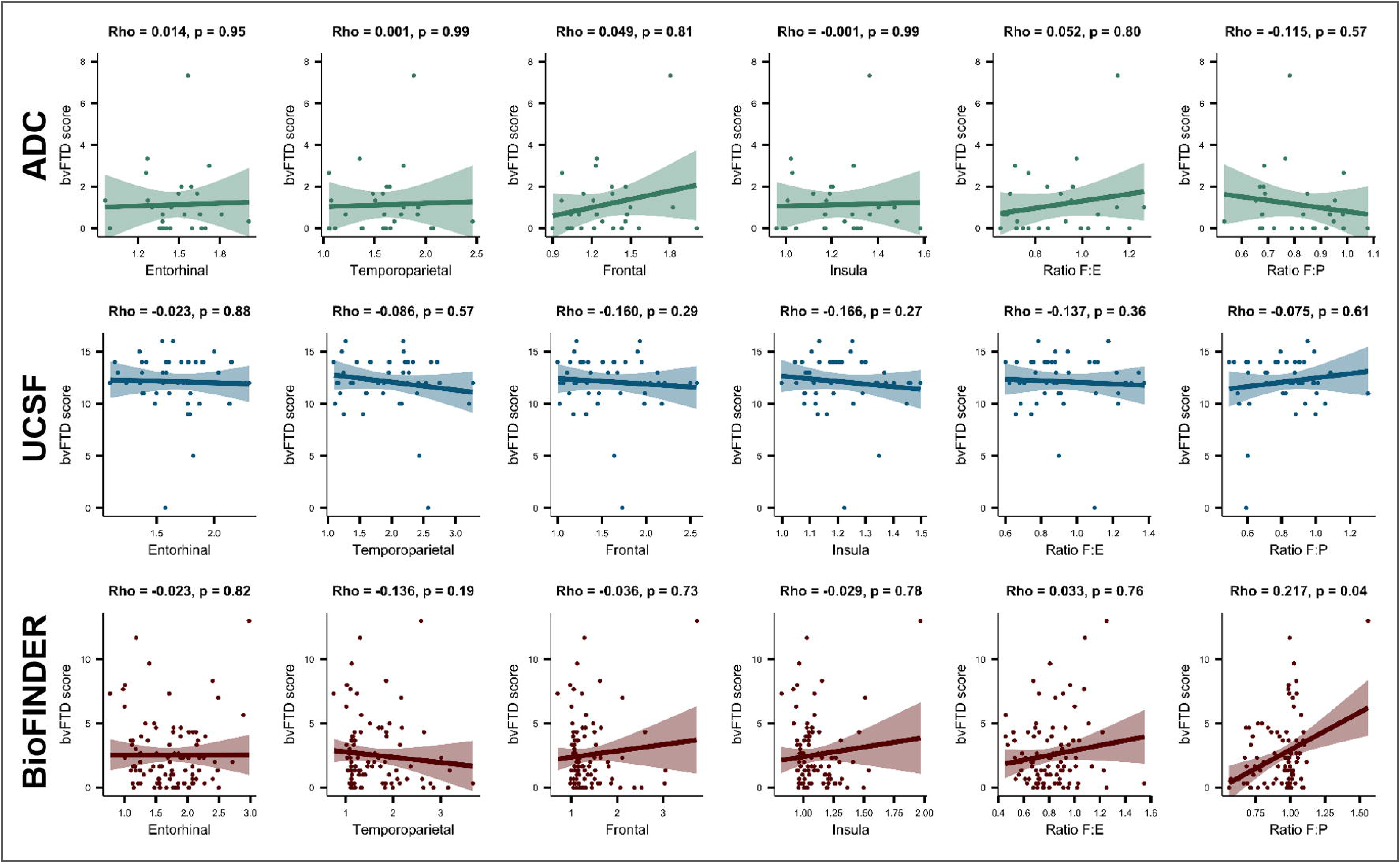
Associations between regional tau PET retention and behavioral variant frontotemporal dementia measures in typical Alzheimer’s disease. Correlations are adjusted for age and sex. *ADC*=Amsterdam Dementia Cohort, *UCSF*=University of California San Francisco, *bvFTD*=behavioral variant of frontotemporal dementia.

## DISCUSSION

In this multicenter case series, we examined the distribution of tau pathology based on PET and postmortem evaluationgs in clinically defined and amyloid-β positive participants with bvAD. In addition, we examined the relationships between tau PET patterns and behavioral symptoms in clinically impaired participants with typical AD. We found a heterogeneous distribution of tau pathology across participants with bvAD, ranging from pronounced anterior involvement to a more temporoparietal pattern based on PET. Immunohistochemistry in an independent sample of bvAD patients supported this heterogeneous distribution of hyperphosphorylated tau pathology across different brain regions, which did not differ from the distribution in typical AD. Finally, we observed that the degree of behavioral impairments was not related to the amount and regional distribution of tau pathology as measured by PET in patients with typical AD. Altogether, these results suggests that tau pathology is not the main driver of the clinical phenotype in bvAD, and that behavioral symptoms (unlike cognition^26^) are not tightly linked to tau pathology in clinically impaired patients with AD.

Our results corroborate the remarkable heterogeneity of tau distributions described in the scarce neuropathological and PET literature on bvAD. While some neuropathological studies indicate more pronounced tau pathology in frontal regions than in other brain regions^6, 27, 28^, others describe a widespread distribution of tau across different lobes in bvAD^13, 29, 30^ participants, or no differences in the burden of frontal tau pathology in bvAD compared to typical AD^4^. However, neuropathological studies typically lack the ability to make inferences on the distributions of tau in early stages of the disease, which is a major advantage of neuroimaging techniques like PET. However, the only *in-vivo* investigations of tau PET in cases with bvAD to date have shown somewhat contradictory results. While one study suggested frontal involvement in addition to a temporoparietal pattern in a bvAD case with advanced dementia (MMSE: 10/30)^13^, another bvAD case with mild dementia (MMSE: 21/30) showed a predominant temporoparietal pattern of tracer retention with sparing of frontal regions^9^. Our extended case series shows that patients with bvAD are primarily characterized by a classical temporoparietal pattern of tau, with, in some cases, pronounced involvement of (mostly lateral) frontal areas, which did not strongly depend on disease severity or age of onset. Importantly, most bvAD cases did not show prominent tau uptake in medial prefrontal and insular regions, which are affected in bvFTD and constitute key regions of the salience network^31^ that regulates complex social behaviors. Indeed, only one case showed disproportionate tau deposition in the medial prefrontal cortex and salience network relative to other brain regions. This is in contrast to other atypical AD variants which almost invariably show tau PET patterns that correspond to their clinical phenotype, i.e. predominant occipito-temporal and/or occipito-parietal involvement in posterior cortical atrophy (the “visual” variant of AD) or highly asymmetric (left > right) tau PET uptake in language network regions in logopenic variant primary progressive aphasia (the “language” variant of AD)^9, 10^. A possible explanation for the discrepancy in bvAD could be that behavioral and socio-emotional processing entail more multifaceted constructs than neurocognitive domains like language and visual functions, and therefore engage wider (sub)cortical regions and networks across the brain^32^.

Besides tau pathology, several other mechanisms may underlie the clinical phenotype in bvAD. First, pathologies other than AD may be driving the behavioral abnormalities. For example, co-occurrence of Lewy body pathology has been observed in more than half of patients with a clinical diagnosis of bvFTD who were neuropathologically diagnosed with AD^33^. However, in our study only one case had a co-primary neuropathological diagnosis of dementia with Lewy bodies in addition to AD, and this low frequency is in accordance with previous pathological findings in clinically defined bvAD^1^. Importantly, no indications for TDP-43 or isolated 3R tau inclusions were found in our bvAD cases and 3/8 cases showed negative TDP-43 staining. As substantial CAA and comorbid Lewy body inclusions were found in both our bvAD and typical AD patients, these comorbid pathologies are likely not driving the differences in clinical phenotypes. Second, patients with bvAD may show lower density of Von Economo Neurons (VENs). VENs are large bipolar projection neurons located exclusively in the anterior cingulate cortex and the frontoinsula^34^ that are affected in bvFTD and psychiatric diseases and are implicated in higher-order social functioning and thus crucial to adaptive behavioral regulation. No significant difference in VEN density was observed in the anterior cingulate cortex between bvAD cases and typical AD cases in a sample of donors with coexisting Lewy body pathology^35^, leaving the role of the VENs in “pure” bvAD unknown. Third, the behavioral disturbances seen in bvAD may arise from damage to deep gray matter or white matter structures that have previously been linked to neuropsychiatric symptoms^36, 37^, rather than from frontal neocortical pathology. However, except for the amygdala, we previously observed no differences in gray matter volumes or patterns of white matter hyperintensities between bvAD and typical AD that are of relevance for behavior^3^. In addition, the current study showed no differences in postmortem tau pathology in subcortical regions between bvAD and typical AD, supporting the notion that these structures may not be disproportionally affected in bvAD. Alternatively, the explanation may lie in functional rather than structural mechanisms, as behavior may rely on complex integrated networks across the brain and we previously showed alterations in metabolic connectivity of the anterior default mode network in bvAD^3^. In addition, analogous to reports of participants with the logopenic variant of progressive aphasia showing learning disabilities in their medical history^38^, the presence of premorbid vulnerable personality structures in participants with bvAD – or a pathological interplay between personality traits and AD pathology^39^ – may provide clues to the clinical phenotype in bvAD. It is conceivable that these vulnerable personality structures are exacerbated once AD pathological changes start to affect the brain, independent of the precise anatomical localization of protein deposition. Future studies should examine this hypothesis, and should also include an assessment of sex differences given the male predominance in bvAD.

The fact that the regional distribution of tau pathology did not show a strong consistent concordance with the clinical features of bvAD raises questions about the relationship between tau and behavior in AD, as bvAD patients represent the extreme of behavioral deficits on the clinical spectrum of AD. Indeed, our investigations of the relationships between regional tau PET uptake and “bvFTD”-like behavioral features suggests that these relationships may be different than those between tau pathology and cognitive functioning in typical^26^ and atypical AD^9, 10^. Although neuropathological studies have found that early neurofibrillary tangle pathology was associated with an increased risk of neuropsychiatric symptoms in AD^40^ and tau PET studies have identified associations between medial temporal tau deposition and a wide range of behavioral measures in preclinical AD^41^, our tau PET study suggests that similar relations are not present in overtly symptomatic phases of AD, and contradict previous reports of associations between frontal regions and behavior in symptomatic AD^42-44^.

Strengths of the current study include the relatively large sample of amyloid-β positive bvAD cases who met ≥2/6 bvFTD criteria and underwent tau PET or autopsy. In addition, comparison to cohort-specific reference groups of typical AD patients aids the clinical interpretation of our findings. Limitations of this study mainly lie in the descriptive nature of in particular the tau PET study, as statistical comparisons were hampered by the small sample size in the bvAD cases due to the low prevalence of this clinical phenotype. In addition, different tau tracers and PET processing pipelines were applied at the different centers, hampering pooling of tau PET data. Second, the presence of comorbid pathology contributing to the clinical presentation cannot be excluded in the tau PET study. Third, the inclusion of right hemispheric regions only in the postmortem evaluations may have created a bias. However, given the demonstrated right hemispheric dominance in bvFTD^45^ and suggested dominance in bvAD^3, 5, 13^ as well as established relationships between right frontal areas and behavioral deficits like apathy, disinhibition and aberrant motor behavior^46^, it is unlikely that this affected our results. Fourth, the comparison of the frontal pole in the postmortem study against the medial and lateral prefrontal cortices in the tau PET study may introduce a bias, as these regions have been differentially implicated in behavioral disturbances^47^. Fifth, although we did not specifically focus on the dysexecutive variant of AD in this study, executive deficits comprised one of the 6 core phenotypic inclusion criteria. Whereas the inclusion of 2/6 bvFTD symptoms strictly allows for inclusion based on one behavioral symptom in addition to executive dysfunction, all cases in our study had at least two behavioral features. Future studies should investigate the differences and overlap between dysexecutive and behavioral variants of AD. Sixth, we only assessed associations between specific “bvFTD” like measures and tau PET uptake, which did not entail the full range of neuropsychiatric symptoms in AD and bvFTD.

Although the neurobiological mechanisms in bvAD are more similar to typical AD than to bvFTD, clinical differentiation between bvAD and bvFTD remains a diagnostic challenge. MRI and [^18^F]FDG-PET provide only modest diagnostic accuracy^1, 3^ and amyloid-β positivity on PET or CSF also occurs frequently in bvFTD patients, especially with advancing age and in the presence of an *APOE* ε4 allele^48^. Tau PET, however, shows very high specificity for tau neurofibrillary tangles in AD dementia^49^, as tau PET signal is low in non-AD neurodegenerative disorders (including sporadic forms of bvFTD^50^). The recent FDA approval for [^18^F]flortaucipir PET for clinical use may therefore aid in the differential diagnosis between bvAD and bvFTD in clinical practice. Ultimately, clinical consensus criteria and standardization of behavioral assessment are necessary to improve diagnosis, prognosis and patient care for individuals with bvAD.

## Data Availability

All data is available upon request.

**Glossary**
AD: Alzheimer’s disease
Aβ: β amyloid
bvAD: behavioral variant of Alzheimer’s disease
tAD: typical Alzheimer’s disease
bvFTD: behavioral variant frontotemporal dementia
MCI: mild cognitive impairment
MMSE: mini mental state examination
APOE: Apolipoproteine E
ADC: Amsterdam Dementia Cohort UCSF –- University of California San Francisco
PET: positron emission tomography

## Notes

### Competing Interest Statement

The authors have declared no competing interest.

### Funding Statement

Work at the Alzheimer Center Amsterdam was supported by the Netherlands Organisation for Health Research and Development, ZonMw (70-73305-98-1214 to Rik Ossenkoppele, PI). Research of the Alzheimer center Amsterdam is part of the neurodegeneration research program of Amsterdam Neuroscience. The Alzheimer Center Amsterdam is supported by Stichting Alzheimer Nederland and Stichting VUmc fonds. Work at the University of California San Francisco was supported by the NIH National Institute on Aging (NIA) grants R01-AG045611 (to G.D.R.) and the Robert W. Katzman Fellowship Training Grant through the American Academy of Neurology in conjunction with the American Brain Foundation and Alzheimer's Association (A133766) to (to W.G.M.), as well as funding for Aging and Dementia Research Center (NIA P30-AG062422) and PPG (NIA P01-AG019724). Work at the Skane University Hospital and Lund University was supported by the Swedish Research Council, the Knut and Alice Wallenberg foundation, the Marianne and Marcus Wallenberg foundation, the Swedish Alzheimer Foundation, the Swedish Brain Foundation, the Skane University Hospital Foundation, and the Swedish federal government under the ALF agreement.

### Author Declarations

Informed consent was obtained from all subjects or their assigned surrogate decision-makers, and the study was approved by the Amsterdam University Medical Center, Memory and Aging Center Clinic at the University of California San Francisco and the Memory Clinic Skane University Hospital institutional human research review boards.

